# A natural experiment in Kenya reveals durable immunosuppressive effects of early childhood malaria: a longitudinal cohort study

**DOI:** 10.1101/2025.05.26.25328345

**Authors:** Mercy S. Safari, Timothy O. Makori, Elijah T. Gicheru, Maureen W. Mburu, Omar Nyawa, Faiz Shee, James Nyagwange, Eunice W. Kagucia, Francis Ndungu, Timothy Chege, James O. Tuju, Charles J. Sande

**Author notes:** Correspondence to: Charles J. Sande, KEMRI-Wellcome Trust Research Programme, Kilifi, Kenya P.O. Box 230 - 80108. Contributed equally (James O. Tuju, Charles J. Sande).

## Abstract

**Background:** Chronic malaria exposure has been proposed to modulate immune function, but its long-term effects on antibody-mediated responses to unrelated pathogens remain poorly defined. Whether these effects persist beyond periods of active infection, and how early-life exposure shapes humoral immunity over time, is not well understood.

**Methods:** We leveraged a natural experiment in coastal Kenya - where two regions (Junju and Ngerenya) diverged sharply in malaria transmission from around 2004 - to evaluate the long-term immunological consequences of malaria exposure in childhood. Using a protein microarray platform, we measured IgG responses to vaccine and pathogen antigens in 123 children sampled longitudinally over a 15-year period. Active weekly malaria surveillance enabled precise reconstruction of individual exposure histories.

**Results:** IgG responses to *Plasmodium falciparum* apical membrane antigen 1 (AMA1) tracked closely with clinical malaria episodes, confirming the ability of the microarray platform to detect biologically meaningful variation in antigen-specific immunity. Despite comparable vaccination histories, children from the high malaria transmission setting (Junju) exhibited persistently lower measles-specific IgG levels than children from the low-transmission setting (Ngerenya), a pattern validated by ELISA. In longitudinal analyses, children from Junju exhibited lower antibody responses to a range of unrelated antigens, including Bordetella pertussis, CMV, rubella, and measles, with similar differences evident in cross sectional analyses at 10 years of age. Within the Ngerenya cohort, children with documented early-life malaria had broadly lower IgG responses at age 10 compared to malaria-naïve peers, despite identical geography, vaccines, and follow-up duration.

**Conclusions:** These findings suggest that malaria exposure during early childhood is linked with durable suppression of antibody responses to unrelated pathogens and vaccines. This effect persists long after infection and may partially explain the overall diminished long-term vaccine effectiveness in malaria-endemic settings.

## Introduction

Malaria remains a major cause of childhood morbidity and mortality in sub-Saharan Africa. In 2023, Africa accounted for 95% of global malaria deaths, with over three-quarters occurring in children under five years old^1^. Alongside malaria, children in these settings face high exposure to a wide array of pathogens - including respiratory, enteric, and helminth infections ^2^, making immune competence critical for survival and long-term health. Despite widespread vaccination efforts, several studies report reduced vaccine efficacy in malaria-endemic regions compared to malaria-naïve populations ^3–5^. Although multiple factors may contribute^6^, growing evidence suggests that malaria itself can impair host immunity^5,7,8^. Repeated P. falciparum infection has been associated with immunomodulatory changes, including expansion of regulatory T cells ^9,10^, regulatory B cells^11^, and atypical memory B cells with limited effector function^12,13^. These alterations promote host tolerance for persistent parasitaemia, resulting from recurrent exposure in endemic settings, but they also suppress effector immune responses^14^, potentially to unrelated antigens, including vaccines. While malaria-induced immunosuppression has been described in both experimental and observational studies^7,8,15,16^, its duration and broader impact on the developing immune system remain poorly understood. Most prior work has focused on short-term or antigen-specific outcomes, leaving open the question of whether early-life malaria exposure durably attenuates antibody responses over the long term. Conflicting findings across settings highlight the need for context-specific, and longitudinal data studies^16,17^.

Here, we address this gap using two longitudinal cohorts from coastal Kenya with contrasting malaria transmission histories. One region (Junju) has experienced a sustained malaria burden, while the other (Ngerenya) underwent a rapid decline in transmission after 2004^18^. Intensive weekly malaria surveillance was conducted over more than a decade, enabling precise classification of individual exposure histories. We combined these data with serial serological measurements using a high-throughput in-house protein microarray. This design enabled us to investigate whether clinical malaria in early life leaves a lasting immunological imprint that compromises antibody responses to common childhood pathogens and vaccines. Our findings reveal a durable suppression of humoral immunity linked to malaria exposure in early childhood, with implications for vaccine effectiveness, serosurveillance interpretation, and immune recovery in endemic regions.

## Materials and Methods

### Study setting and cohort design

This study was conducted in Kilifi County, a rural region on the northern coast of Kenya, within the catchment area of the Kilifi Health and Demographic Surveillance System (KHDSS), a long-term population-based platform maintained by the KEMRI-Wellcome Trust Research Programme^19^. Serum samples were obtained retrospectively from two well-characterised paediatric cohorts - Ngerenya and Junju - enrolled between 1998 and 2017 as part of annual malaria cross-sectional surveys^20^. Ngerenya and Junju were selected for their contrasting malaria transmission intensities; Ngerenya experienced a sharp decline in malaria transmission beginning in the early 2000s^18^, whereas Junju maintained moderate malaria endemicity throughout the study period, with *P.falciparum* prevalence approximating 30% during the rainy seasons^21^. All children were visited weekly at home for the detection of febrile episodes, and any child with an axillary temperature ≥37.5°C was tested for P. falciparum parasitaemia using a rapid diagnostic test, with confirmation by microscopic examination of Giemsa-stained thick and thin blood smears. A clinical malaria episode was defined as fever in the presence of ≥2,500 parasites/μL. In addition to active malaria surveillance, serum samples were collected annually from each child and for future serological analysis. The vaccination status of each child for routine childhood vaccines was assessed using digitised immunisation records stored at the KEMRI-Wellcome Trust Research Programme.

### Protein microarray antibody profiling

Antibody responses were measured using an in-house protein microarray platform. Recombinant and whole-virus lysate antigens (Supplementary table 1) were reconstituted in a glycerol-based buffer containing 1% Triton X-100 and printed in duplicate onto epoxy-coated glass slides using a non-contact microarrayer (Marathon Argus, Arrayjet, Scotland). In addition to antigen spots, each miniarray contained a series of internal controls. These included anti-human IgG and anti-human IgA capture antibodies to confirm the presence and isotype of immunoglobulin in each sample, fluorophore-conjugated IgG and IgA (Alexa Fluor 647 and Alexa Fluor 555) to assess scanner performance independently of antigen binding, and printing buffer-only spots to quantify non-specific background signal. Slides were fitted into hybridisation cassettes, washed with PBST (0.05% Tween-20 in PBS), and blocked for 1 hour at 37°C using PBST containing 5% BSA. Serum samples were diluted 1:30 in PBST with 5% BSA and incubated on the slides for 3 hours at room temperature. For each slide, one miniarray was incubated with PBS in place of serum as a negative control, and one miniarray with pooled adult serum, comprising sera from multiple healthy adults, to provide a consistent positive reference for antigen recognition across slides. Following incubation, slides were washed and probed with secondary antibodies: goat anti-human IgG conjugated to Alexa Fluor 647 and goat anti-human IgA conjugated to Alexa Fluor 555, enabling simultaneous detection of IgG and IgA binding. Slides were scanned using a GenePix 4300A scanner with dual-wavelength acquisition (635 nm and 532 nm) to capture isotype-specific signals. To improve measurement robustness and account for spatial variation, each sample was assayed on two independent miniarrays per slide, yielding four spatially separated replicate measurements per antigen. Mean fluorescence intensities (MFIs) were extracted and background-corrected using printing buffer and negative control spots to account for non-specific signal. Technical variation was assessed by calculating the coefficient of variation (CV) across the four replicate spots for each antigen. Measurements with CV >20% were excluded, and retained values were averaged to generate a single antigen-specific response per sample. Pooled adult serum controls were used to monitor inter-slide consistency over time. All data processing and quality control steps were implemented in R (version 4.4.2).

### Enzyme-linked immunosorbent assay (ELISA)

Measles-specific IgG was quantified using a conventional direct ELISA. Plates were coated overnight at 4°C with 2.30 µg/mL measles antigen diluted in PBS. After blocking with 5% skimmed milk for 1 hour at 37°C, serum samples (1:100 dilution) were added and incubated for 1.5 hours at 37°C. Plates were washed with PBST and incubated with HRP-conjugated secondary antibody (1:100 dilution) for 1 hour. Following additional washes, 100 µL of OPD substrate solution (30 mg OPD in 30 mL PBS with 30 µL H₂O₂) was added and incubated in the dark for 10 minutes. Reactions were stopped with 50 µL of 2.5 M sulfuric acid, and absorbance was measured at 490 nm.

### Statistical analysis

All statistical analyses were performed using R (version 4.4.2). Antibody responses between cohorts were compared using the Wilcoxon rank-sum test. P-values <0.05 were considered statistically significant. To compare antibody responses between the Junju and Ngerenya cohorts, longitudinal analyses were performed using linear mixed-effects models, which accommodate unbalanced data and allow inclusion of all available observations without requiring imputation. Antibody responses were modelled as a function of cohort, with age included as a non-linear term and a random intercept for each child to account for repeated measurements. From these models, we estimated the average age-adjusted difference in antibody responses between cohorts across the full follow-up period. P-values were adjusted for multiple antigen testing using the false discovery rate (FDR) method. To examine the relationship between malaria exposure and heterologous antibody responses, we used cumulative febrile malaria episode count derived from longitudinal surveillance data as a measure of long term exposure. Antibody measurements were log transformed prior to analysis, and values for each antigen were standardised to z scores to enable comparison of responses to different antigens with differing dynamic ranges. Associations between malaria exposure and antibody responses were assessed using linear mixed-effects models, with malaria episode count as the primary exposure, age modelled as a non-linear term, and random intercepts for both child and antigen to account for repeated measurements and between-antigen variability. For visualisation, unadjusted scatterplots with fitted linear trends were used to illustrate the relationship between malaria episode burden and antibody responses, stratified by antigen. To assess potential population-level differences in nutritional status between regions, we analysed contemporaneous hospital-based surveillance data from the same geographic populations. Anthropometric measures (mid-upper arm circumference (muac), weight-for-age, and height-for-age) were modelled using linear mixed-effects regression, with location (Junju vs Ngerenya) as the primary exposure. Age and calendar year were modelled using natural cubic splines to account for non-linear effects, and models were adjusted for concurrent infections (RSV, parainfluenza, influenza A, human metapneumovirus, OC43, and malaria). Data were anonymised and delinked from all personally identifiable information.

### Ethics statement

The study was approved by the Scientific and Ethics Review Unit (SERU) of the Kenya Medical Research Institute. All procedures were conducted in accordance with the principles of Good Clinical Laboratory Practice (GCLP).

## Results

### A natural experiment in coastal Kenya reveals sharply divergent malaria exposure trajectories in early childhood

Between 1998 and 2017, two rural communities in coastal Kenya - Junju and Ngerenya - underwent markedly different transitions in malaria transmission. Both regions experienced a high malaria burden in the 1990s and early 2000s. However, beginning in 2004, transmission in Ngerenya declined sharply and remained near zero for more than a decade, while Junju continued to experience sustained endemicity well into the mid-2010s. To quantify the scale and timing of this transition, we analysed surveillance data collected from August 1998 to April 2017. A total of 1,243 children were followed in Ngerenya and 659 in Junju. In Ngerenya, the proportion of febrile surveillance visits with confirmed P. falciparum parasitaemia declined from 22.4% before 2004 (1,378 of 6,148 visits) to just 1.1% after 2004 (48 of 4,389 visits). In contrast, Junju children continued to experience high rates of febrile malaria, with 17% of visits testing positive between 2007 and 2017 (869 of 5,130 visits) - **Fig. 1**. Baseline cohort characteristics are shown in **Table 1**.

**Figure 1.**
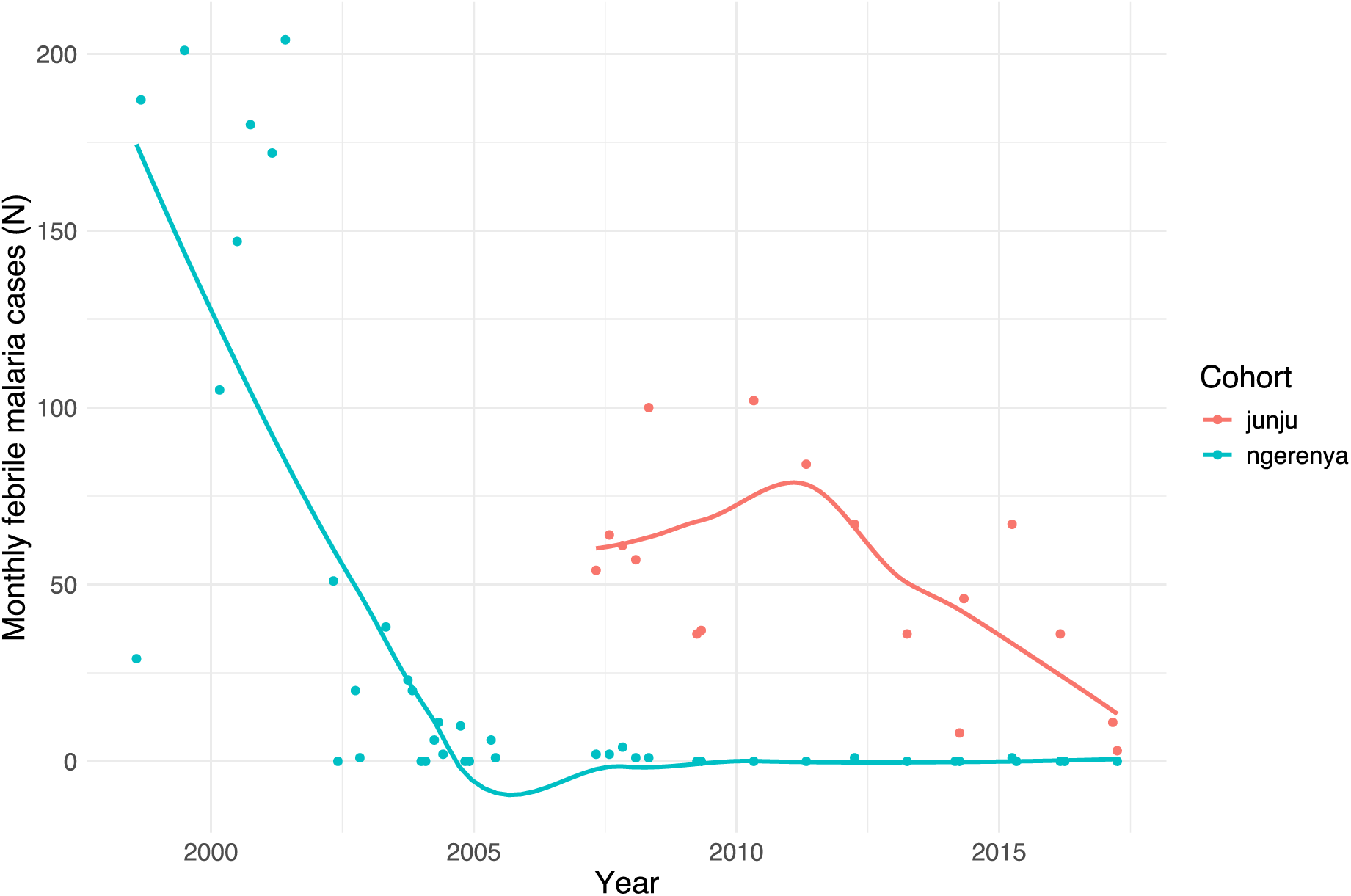
Divergent malaria exposure histories in coastal Kenya. Monthly malaria case counts from active surveillance in two adjacent regions of Kilifi County, Kenya, between 1998 and 2017. Junju (red) maintained sustained malaria transmission throughout the study period, while Ngerenya (blue) experienced a rapid collapse in transmission beginning in 2004. Points represent total cases per month; lines show smoothed trends generated using locally weighted regression (loess) in R (span = 0.8).

**Table 1:**
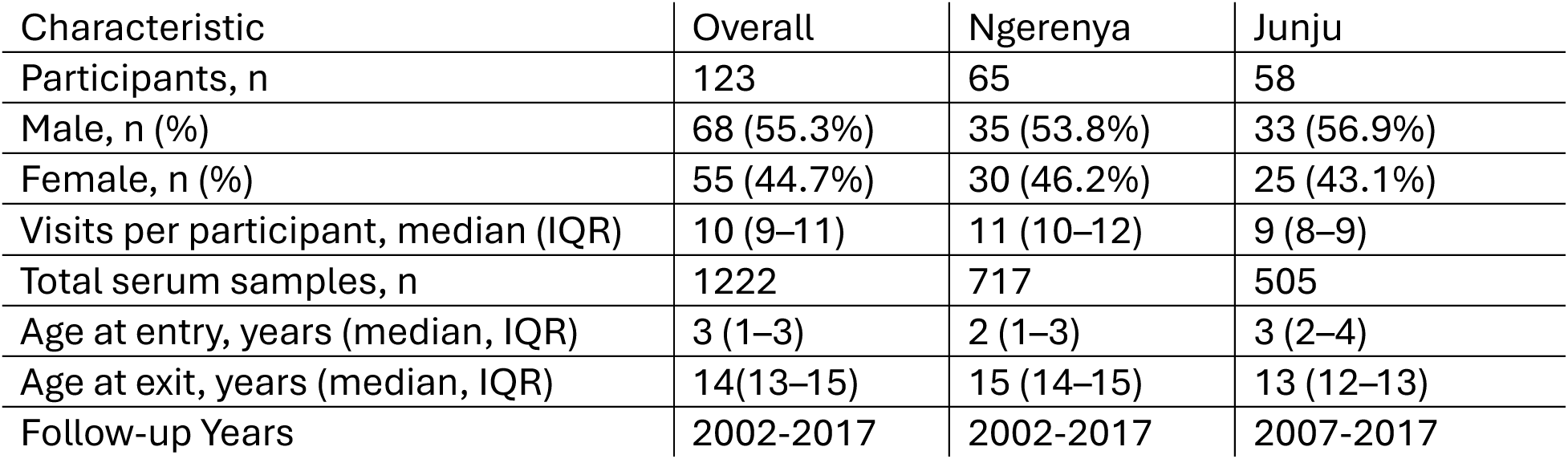
Baseline characteristics of the longitudinal cohorts from Junju and Ngerenya.

| Characteristic | Overall | Ngerenya | Junju |
| --- | --- | --- | --- |
| Participants, n | 123 | 65 | 58 |
| Male, n (%) | 68 (55.3%) | 35 (53.8%) | 33 (56.9%) |
| Female, n (%) | 55 (44.7%) | 30 (46.2%) | 25 (43.1%) |
| Visits per participant, median (IQR) | 10 (9–11) | 11 (10–12) | 9 (8–9) |
| Total serum samples, n | 1222 | 717 | 505 |
| Age at entry, years (median, IQR) | 3 (1–3) | 2 (1–3) | 3 (2–4) |
| Age at exit, years (median, IQR) | 14(13–15) | 15 (14–15) | 13 (12–13) |
| Follow-up Years | 2002-2017 | 2002-2017 | 2007-2017 |

To assess whether the protein microarray platform accurately captured longitudinal P. falciparum-specific immune responses, we examined IgG levels against the plasmodium apical membrane antigen 1 (AMA1) in individual children with known malaria exposure histories. Among children with multiple confirmed episodes of febrile malaria detected via weekly active surveillance, AMA1-specific IgG levels increased sharply over time, tracking closely with the timing of clinical infections (example shown in **Fig. 2a**), while children from Ngerenya who remained malaria-free throughout follow-up exhibited consistently low IgG levels against AMA1 over more than a decade of surveillance (example shown in **Fig. 2b**). These patterns mirrored expected immunological trajectories of repeated exposure versus non-exposure, confirming that the microarray platform is capable of detecting meaningful variation in P. falciparum-specific humoral responses. We then extended this analysis to assess longitudinal IgG responses to AMA1 in a subset of 123 children drawn from the two surveillance cohorts. These children contributed a total of 1222 serum samples, with a median of 10 samples per child, collected between 2002 and 2017 (**Supplementary Fig. 1**). This subset was selected on the basis of the relative completeness of their longitudinal follow-up serum sampling and included 58 children from Junju (505 samples) and 65 from Ngerenya (717 samples). AMA1-specific IgG levels diverged sharply between cohorts early in life and remained distinct throughout follow-up. In Ngerenya, levels declined rapidly after 2003, stabilising at lower levels by mid-childhood. In contrast, Junju children maintained elevated levels across all time points (**Fig. 2c**).

**Figure 2.**
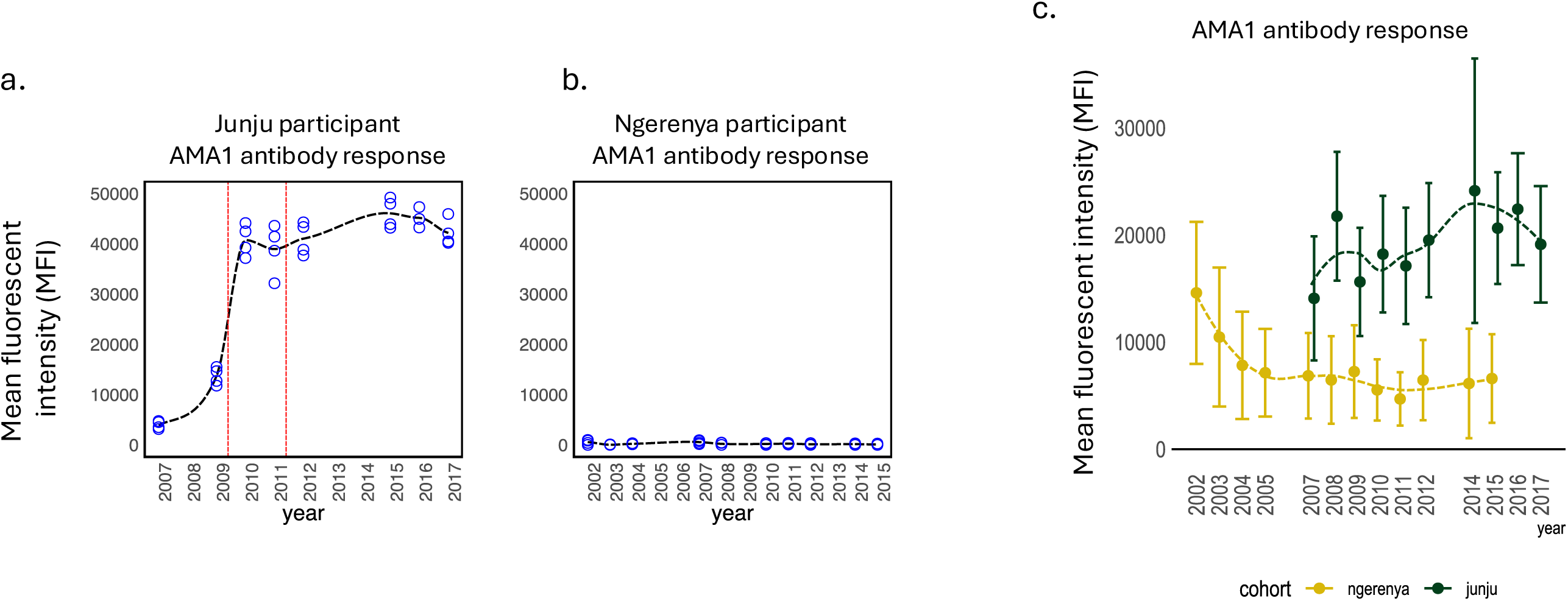
AMA1-specific IgG trajectories mirror individual and regional malaria exposure. **(A–B)** Longitudinal AMA1 IgG levels in individual children measured by protein microarray. Vertical red lines denote confirmed febrile malaria episodes. Panel A shows a Junju child with multiple documented infections; Panel B shows a Ngerenya child who remained malaria-free throughout follow-up. Each blue spot is a single antibody measurement. Each time point was measured in quadruplicate **(C)** Mean AMA1-specific IgG levels with 95% confidence intervals for all children in the microarray subset plotted by year of sampling. Junju children showed persistently elevated antibodies, while AMA1 antibody levels in Ngerenya declined sharply after 2004.

### Malaria-exposed children exhibit lower antibody levels to non-malarial antigens

To examine the impact of differential malaria endemicity on the antibody response to non-malarial antigens, we first compared IgG responses to the measles virus among Junju and Ngerenya children. We started by validating the ability of the in-house protein microarray platform to detect biologically meaningful measles-specific IgG responses by examining longitudinal measles levels in individual children with known vaccination histories. Among children with a documented history of receiving all three recommended doses of measles vaccine, we observed a sharp rise in IgG levels following immunisation, followed by sustained levels into later childhood (example shown in **Fig. 3a**). In contrast, unvaccinated children exhibited consistently low IgG trajectories across all time points (example shown in **Fig. 3b**). These patterns were reproducible across the cohort and recapitulated expected vaccine-induced versus naïve antibody dynamics, supporting the use of this platform for population-level serological inference. Using this platform, we then compared IgG responses to measles virus among children from Junju and Ngerenya. This analysis was restricted only to children with a documented record of measles vaccination. Despite matched vaccination histories, children from Junju - where malaria transmission remained high - consistently exhibited lower measles-specific IgG titres than children from Ngerenya, where malaria transmission had declined. This difference was evident from the earliest timepoints and persisted throughout childhood. Annual antibody measurements showed that mean measles-specific IgG levels were higher in Ngerenya than in Junju in every sampling year (**Fig. 3c**). To validate these findings, we measured measles-specific IgG levels by ELISA in a subset of 3-year-old children who had completed measles vaccination at least one year prior. The assay included a negative control (PBS) and pooled adult serum as a positive reference. Consistent with the microarray results, Junju children again displayed significantly lower IgG levels than their Ngerenya counterparts (**Fig. 3d**).

**Figure 3.**
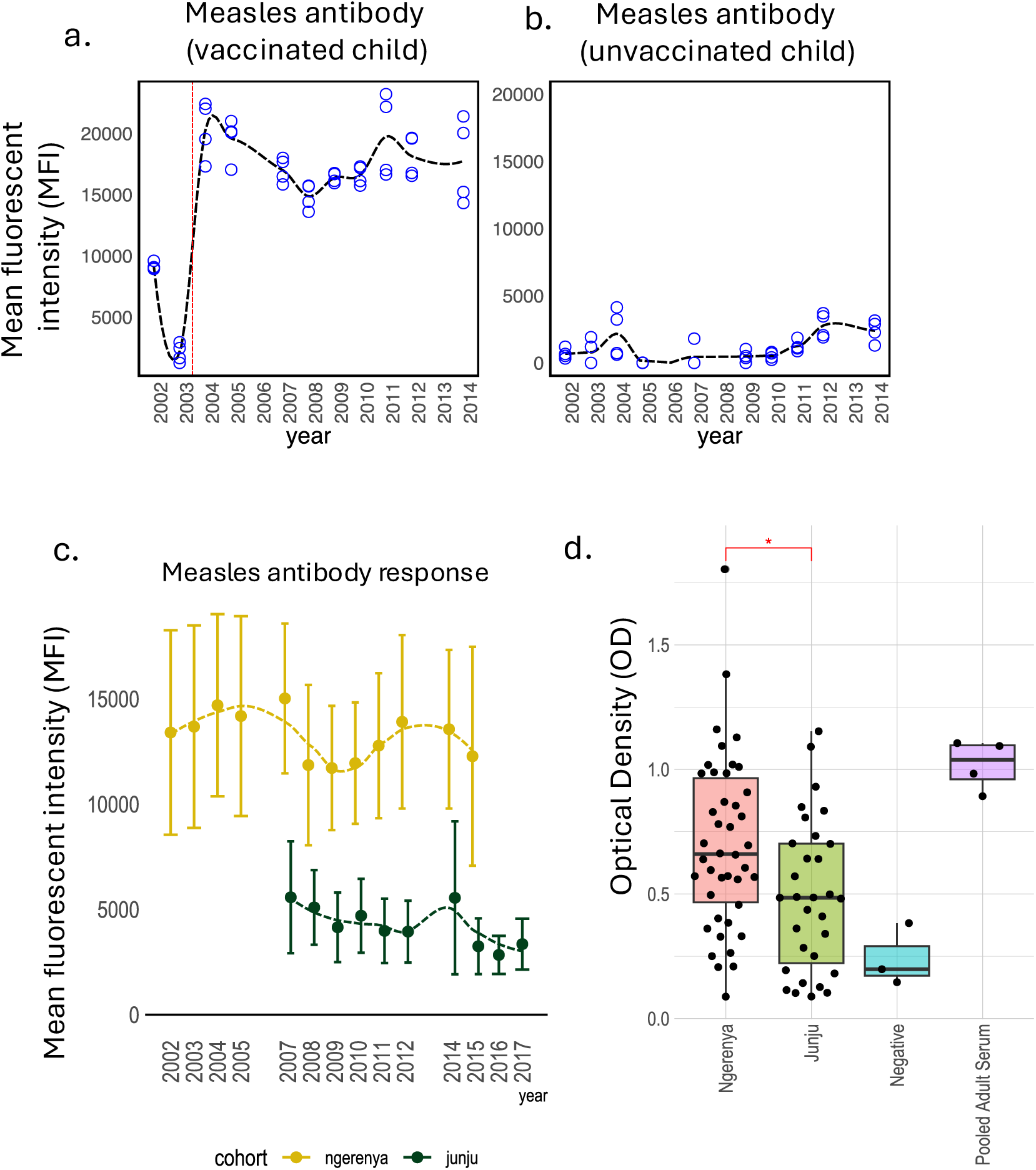
Malaria exposure is associated with reduced measles-specific antibody levels. Example plots of temporal changes in measles-specific antibody in vaccinated and unvaccinated children are shown **(A)** Longitudinal IgG levels in an individual child measured by microarray. The dashed vertical red line indicates the timing of the last dose of the routine measles vaccine. **(B)** Shows a similar temporal trend for a child with no history of measles vaccination. **(C)** Longitudinal IgG responses to measles virus by cohort, measured by protein microarray in vaccinated children. Junju children exhibited consistently lower levels of measles-specific antibody than Ngerenya counterparts. The circles indicate means, and the whiskers denote 95% confidence intervals. **(D)** Measles-specific IgG levels in 3-year-old children from Junju and Ngerenya, measured by ELISA in children with a documented history of measles vaccination, including a negative control (PBS) and pooled adult serum as a positive reference Each dot represents an individual participant.

### Early-life malaria exposure is associated with long-term suppression of antibody responses

To assess whether the attenuation of antibody responses extended beyond measles, we compared IgG levels to a broader panel of antigens, including vaccine-preventable pathogens (*Bordetella pertussis*, H1N1 influenza virus, rubella virus, and measles virus) and common childhood infections (cytomegalovirus (CMV), Epstein–Barr virus (EBV), herpes simplex virus 1 (HSV-1), and coxsackievirus B1). Using mixed-effects models incorporating all available longitudinal measurements, children from Ngerenya exhibited higher antibody responses than those from Junju after adjustment for age and repeated measurements within individuals (**Fig. 4a,b**). Effect estimates were consistent in direction across most antigens, with particularly marked differences for HSV-1, EBV and measles. Differences were also observed for coxsackievirus B1 and *Bordetella pertussis*, while smaller or non-significant differences were seen for CMV, rubella, and H1N1 influenza. Several of these associations remained statistically signifcant after correction for multiple testing (**Fig 4b**). To complement these longitudinal analyses, we performed cross-sectional comparisons of antibody responses at 10 years of age. This showed a similar pattern, with children from Junju exhibiting lower IgG levels for most antigens compared to their Ngerenya counterparts (**Fig. 4c**). Differences were particularly marked for coxsackievirus, EBV, HSV-1, and measles. Antibody responses to the 2009 pandemic strain of H1N1 were the least differentiated between cohorts.

**Figure 4.**
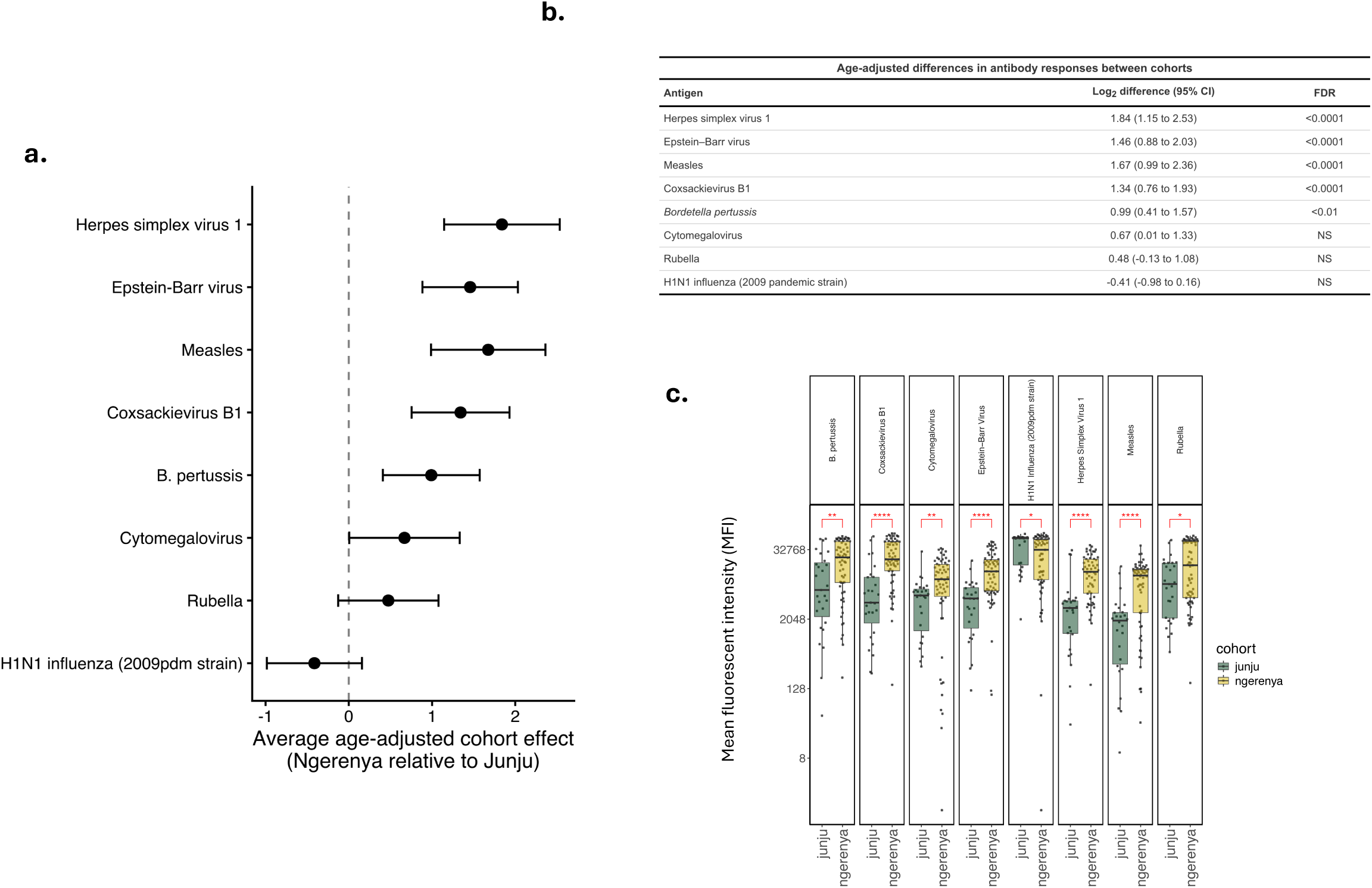
Early-life malaria exposure is associated with reduced antibody responses to multiple antigens. **(A)** Forest plot showing the average age-adjusted difference in log₂ antibody responses between children from Ngerenya and Junju, estimated using mixed-effects models incorporating all available longitudinal measurements. Points represent model estimates and horizontal bars indicate 95% confidence intervals. **(B)** Summary table of model-derived estimates, including log₂ differences with 95% confidence intervals and false discovery rate (FDR)-adjusted significance across antigens. **(C)** Cross-sectional comparison of antibody responses at 10 years of age, shown for reference. Boxes indicate interquartile ranges, centre lines denote medians, and whiskers represent 1.5× the interquartile range. Asterisks indicate significance from Wilcoxon rank-sum tests (* P < 0.05, ** P < 0.01, *** P < 0.001, ****. P < 0.0001).

To determine whether the attenuation of antibody responses could still be attributed to early-life malaria exposure independent of geographic or environmental differences, we conducted a stratified analysis within the Ngerenya cohort, which experienced a sharp decline in malaria transmission beginning in 2004. Due to stochastic differences in malaria infection around the time of this inflection, children in Ngerenya had different malaria exposure histories despite living in the same area. At the 10-year-of-age time point, sera were available for 62 out of the 65 children that were originally selected in the Ngerenya cohort subset for serological analysis. Of these, 20 experienced one or more episodes of febrile malaria during early childhood prior to the decline in malaria transmission, while 42 remained entirely malaria-free throughout follow-up (**Fig. 5a**). At 10 years of age, Ngerenya children who had experienced early-life malaria exhibited significantly lower IgG levels to a wide range of antigens compared to their malaria-naïve peers (**Fig. 5b**). These included responses to Coxsackievirus B1, CMV, H1N1 influenza, HSV-1 and rubella. Differences in antibody level to *B. pertussis* and EBV did not reach statistical significance.

**Figure 5.**
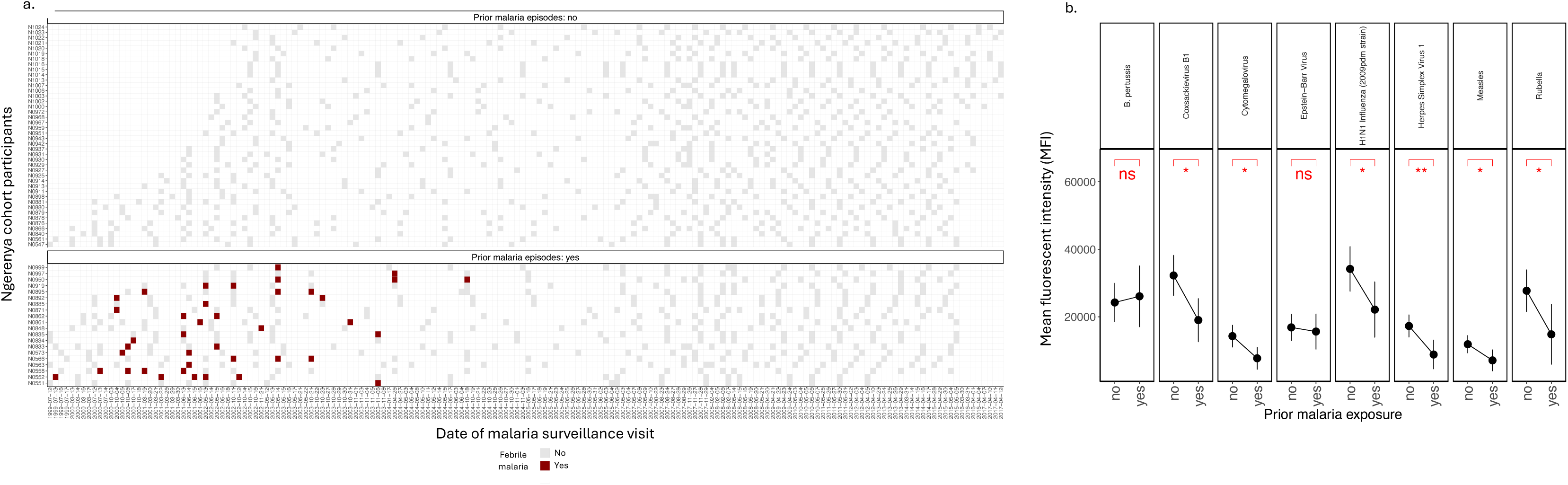
Early-life malaria exposure predicts long-term suppression of antibody responses within the same geographic region. **(A)** Active malaria surveillance records for children in the Ngerenya cohort. Each row represents an individual child, and each column represents a surveillance timepoint. Dark red boxes indicate one or more confirmed febrile malaria episodes; light grey boxes indicate surveillance visits without malaria detection. Children are grouped by early-life exposure status (top: malaria-naïve; bottom: previously exposed). **(B)** IgG levels at 10 years of age among Ngerenya children, stratified by early-life malaria exposure. Children with ≥1 confirmed febrile malaria episode during early childhood (n = 20) show significantly lower titres to multiple unrelated pathogens compared to malaria-naïve peers (n = 42). All children lived in the same geographic area and received identical vaccines and follow-up. The black dots are means and error bars are 95% confidence intervals.

### Population-level comparison of anthropometric and infection profiles between regions

Because anthropometric measurements were not collected routinely within the longitudinal malaria cohorts, we assessed potential population-level differences in nutritional status using contemporaneous hospital-based surveillance data from the same geographic regions. This dataset comprised repeated measurements of mid-upper arm circumference (MUAC), weight-for-age, and height-for-age across early childhood, alongside virological and malaria diagnostics, providing an independent view of the underlying populations from which the longitudinal cohort was drawn. Across early childhood, age-specific distributions of MUAC, weight-for-age, and height-for-age were broadly similar between children from Junju and Ngerenya, with overlapping distributions at all ages (**Fig. 6a**). To formally assess these differences, we fitted regression models adjusting for age, calendar year, and concurrent infections (RSV, parainfluenza, influenza A, human metapneumovirus, OC43, and malaria). Across all three anthropometric indices, there was no evidence of systematic differences between the two populations (**Fig. 6b,c**). Adjusted differences between Junju and Ngerenya were small and centred around zero (MUAC: −0.12, 95% CI −0.38 to 0.15; weight-for-age: −0.05, −0.28 to 0.19; height-for-age: 0.08, −0.17 to 0.33). Notably, effect estimates were small and confidence intervals spanned zero in all cases.

**Figure 6.**
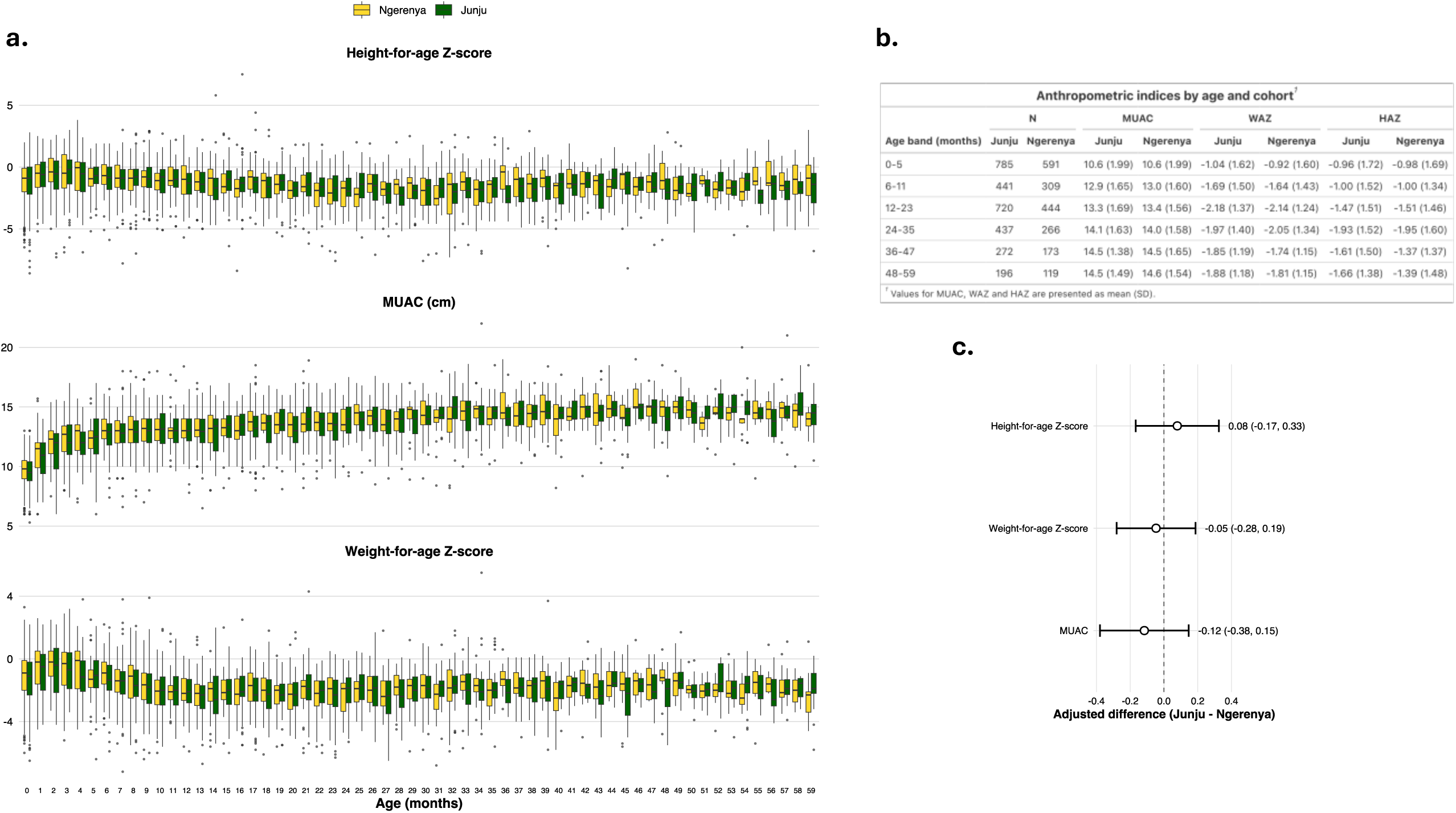
Population-level comparison of anthropometric profiles between Junju and Ngerenya. **(A)** Age-specific distributions of height-for-age (HAZ), mid-upper arm circumference (MUAC), and weight-for-age (WAZ) among children aged 0–59 months, derived from contemporaneous hospital-based surveillance data. Boxplots show median and interquartile range, with whiskers extending to 1.5× the interquartile range; points represent individual observations. Distributions are shown separately for children from Junju and Ngerenya. **(B)** Summary of anthropometric indices by age band and location. Values are presented as mean (standard deviation) for MUAC and mean (95% confidence interval) for WAZ and HAZ. **(C)** Adjusted differences in anthropometric indices between children from Junju and Ngerenya. Points represent model-derived estimates for the difference (Junju − Ngerenya), and horizontal lines indicate 95% confidence intervals. Estimates were obtained from regression models adjusting for age (modelled using splines), calendar year, and concurrent infections (RSV, parainfluenza, influenza A, human metapneumovirus, OC43, and malaria)

### Higher malaria episode burden is associated with a graded reduction in heterologous antibody responses

To further examine whether the attenuation of antibody responses varied in relation to the intensity of malaria exposure, we analysed the association between cumulative febrile malaria episode count and antibody responses in the longitudinal cohort. Antibody responses were standardised within antigen to enable comparison for the panel of heterologous antigens. In mixed-effects models incorporating all available longitudinal measurements and adjusting for age and repeated measures, higher malaria episode burden was associated with lower heterologous antibody responses (β = −0.086, 95% CI −0.142 to −0.029, p = 0.003). When examined separately by antigen, the direction of association was consistent for the majority of antigens, including Bordetella pertussis, coxsackievirus B1, Epstein–Barr virus, herpes simplex virus 1, measles virus, and rubella virus, with no evidence of opposing trends. Associations for cytomegalovirus and H1N1 influenza were weaker, but did not contradict the overall pattern (**Fig. 7**).

**Figure 7.**
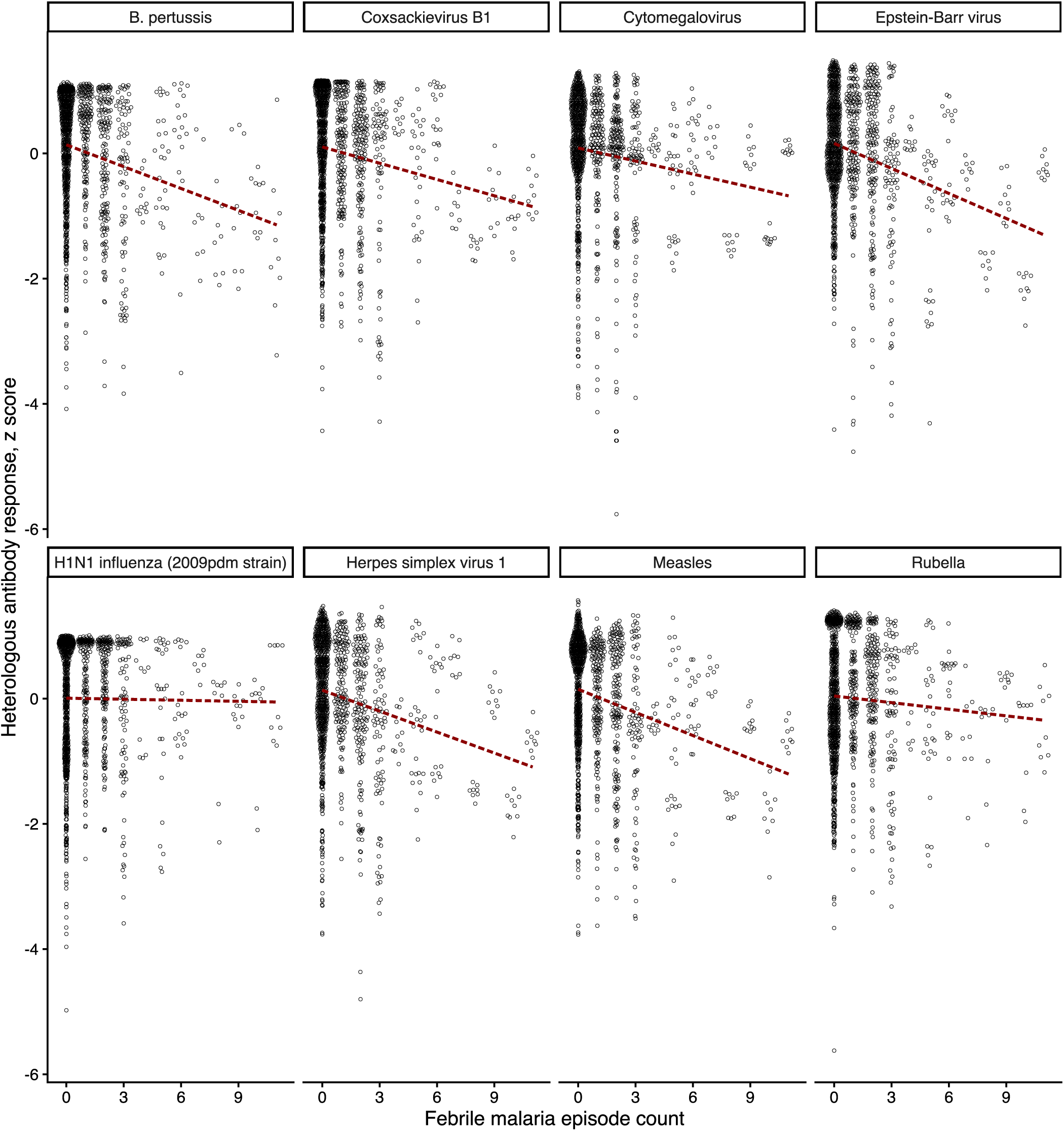
Febrile malaria episode burden is inversely associated with heterologous antibody responses. Scatterplots show the relationship between cumulative febrile malaria episode count and standardised antibody responses (z scores) to eight heterologous antigens. Each point represents an individual observation, and dashed lines indicate fitted linear trends. For most antigens, higher malaria episode burden was associated with lower antibody responses, with no evidence of opposing trends.

## Discussion

This study demonstrates that early-life exposure to malaria is associated with broad and durable impairments in antibody-mediated immunity to unrelated pathogens and vaccines. By leveraging a natural experiment in coastal Kenya - where malaria transmission diverged sharply between adjacent communities during the early 2000s - we were able to disentangle the immunological effects of malaria exposure from confounding geographic and temporal factors. Our findings reveal that children exposed to malaria in early childhood not only generate lower antibody titres to non-malarial antigens but maintain these attenuated responses well into adolescence, long after malaria transmission has ceased. The attenuating effect of malaria on antibody responses has long been suspected ^15^ but difficult to quantify. Prior studies have shown reduced vaccine efficacy in malaria-endemic settings ^3,16^, and experimental models have implicated regulatory T cells, atypical memory B cells, and B cell exhaustion as potential mediators of malaria-induced immune suppression^9,12,13,22^. However, most human studies to date have focused on short-term immune responses or outcomes in the setting of concurrent parasitaemia. Our data extend this work by demonstrating that transient exposure to malaria in early childhood is sufficient to imprint long-lasting changes on the humoral immune repertoire.

The strength of this study lies in its integration of long-term active malaria surveillance with serial antibody profiling. Children were visited weekly for febrile illness surveillance, allowing for precise documentation of clinical malaria episodes. In parallel, our use of a validated protein microarray platform enabled us to track IgG responses to a wide panel of pathogen and vaccine antigens at multiple timepoints. We first confirmed that the microarray platform captured biologically relevant responses by comparing antibody trajectories in individual children with known measles vaccination and malaria exposure histories. The platform reliably detected both vaccine-induced and infection-associated rises in IgG, providing confidence in the longitudinal patterns observed. We found that children from Junju, a region of persistent transmission, had significantly lower antibody levels to multiple pathogens compared to children from Ngerenya, where malaria transmission declined in the mid-2000s. To ensure that these differences were not driven by cross-sectional comparisons at a single timepoint, we additionally analysed antibody responses using mixed-effects models incorporating all available longitudinal measurements. Across multiple antigens, children from Ngerenya exhibited higher antibody responses than those from Junju after adjustment for age and repeated measurements within individuals. Effect estimates were consistent in direction across most antigens and remained evident after correction for multiple testing, indicating that the observed differences reflect a generalised, age-adjusted cohort effect rather than a feature of a specific age or sampling timepoint. Importantly, all children had comparable vaccination histories and were followed through the same longitudinal infrastructure, minimising the likelihood of differential healthcare access or vaccine uptake.

To test whether early-life malaria exposure specifically contributed to long-term immune suppression, we examined antibody responses within the Ngerenya cohort. Because the transmission decline occurred rapidly, children born just before and after the inflection point experienced markedly different levels of malaria exposure while living in the same geographic area. Among children followed longitudinally, those with even limited early-life exposure to malaria had significantly lower antibody titres at 10 years of age than their malaria-naïve peers. This within-cohort contrast strongly implicates early-life infection as the critical window for immune programming. Taken together, these findings support a model in which malaria exposure during critical developmental windows modulates long-term maintenance of immune memory. This may occur through direct effects on B cell maturation, altered antigen presentation^23^, or long-lived changes in lymphoid microenvironments^24–26^. While this study was not designed to resolve the mechanistic basis of these observations, the pattern we describe is consistent with a growing body of evidence highlighted above, that malaria infection can induce sustained perturbations in both B cell and T cell compartments. These studies have demonstrated expansion of atypical memory B cells, disruption of germinal centre responses, and increased regulatory immune activity following malaria exposure, all of which may impair the generation and maintenance of effective humoral immunity. In this context, our findings provide population-level evidence of a durable alteration in antibody profiles associated with early-life malaria exposure. The extent to which these changes reflect persistent alterations in immune cell function or the cumulative effects of repeated infection remains to be determined and will require targeted mechanistic studies. Consistent with this, analyses using cumulative febrile malaria episode count demonstrated a graded inverse association between malaria burden and antibody responses across a broad panel of heterologous antigens, with effects evident across childhood. This supports a dose-dependent relationship between malaria exposure and long-term attenuation of humoral immunity.

This work has important implications for vaccine policy and infection risk in malaria-endemic regions. Reduced antibody titres to vaccine-preventable diseases may translate into diminished long-term protection, even when vaccine coverage is high. Our findings raise the possibility that children in high-transmission settings may require different immunisation strategies, such as delayed dosing and boosting, to improve vaccine-induced immunity. Moreover, as malaria control efforts continue to shift disease epidemiology, these data highlight the value of longitudinal serological surveillance for understanding the broader immunological legacy of malaria exposure. Our study has several limitations. Although the microarray platform was validated internally and against ELISA, quantitative comparisons across platforms are inherently constrained.

Additionally, while the sample size for antibody profiling was modest, the inclusion of dense longitudinal sampling with weekly malaria surveillance and repeated serological measurements provides high-resolution insight into exposure and immune response trends, and the consistency of findings across multiple antigens, analytical approaches, and assay platforms supports the robustness of the observed effects. Finally, we cannot fully exclude the influence of other infections or environmental exposures that may have differed subtly between subgroups, although the within-Ngerenya analysis provides strong evidence for malaria as a primary driver of long-term immune attenuation. Because the longitudinal cohort was originally designed to characterise the acquisition of naturally acquired immunity to malaria, anthropometric measurements were not collected systematically within that dataset, precluding direct adjustment for nutritional status in the primary analyses. To address this, we analysed contemporaneous hospital-based surveillance data from the same geographic regions, comprising measurements of anthropometry and infection status in early childhood. Across three independent indices of nutritional status (MUAC, weight-for-age, and height-for-age), we found no evidence of systematic differences between children from Junju and Ngerenya after adjustment for age, calendar year, and concurrent infections. Effect estimates were small, crossed zero. As the longitudinal cohorts in Junju and Ngerenya were drawn from these underlying populations, these findings suggest that the two groups were broadly comparable with respect to early-life growth and nutritional status, and make it unlikely that nutritional differences are a major driver of the observed immunological patterns.

While this study demonstrates consistent and durable differences in antibody levels across a wide range of antigens, it does not include functional immunological assays to determine the downstream consequences of these differences. As such, we are unable to directly assess whether the lower antibody titres observed in malaria-exposed children translate into reduced neutralising capacity or diminished clinical protection. The primary aim of this study was to identify long-term alterations in humoral immune profiles associated with early-life malaria exposure, rather than to resolve their functional significance. Future studies incorporating functional assays and clinical outcome data will be important to determine the extent to which these serological differences translate into altered susceptibility to infection.

In summary, our findings reveal that early-life malaria exposure is associated with long-term suppression of antibody responses to unrelated pathogens and vaccines. This effect is detectable many years after infection and appears to persist even in the absence of ongoing transmission. As global malaria control efforts continue, understanding the immunological legacy of childhood malaria may be critical for improving vaccination strategies and mitigating susceptibility to other infections.

## Data and Code Availability

The datasets and R scripts used for the analysis in this study are available from the corresponding author upon reasonable request. Due to local laws and regulations, individual-level data from the Kilifi Health and Demographic Surveillance System (KHDSS), cannot be made publicly available. All data processing and visualisation were performed using open-source R packages; package versions and analysis pipelines are available upon request.

## Funding statement

This study was supported by fellowship funding to C.J.S. from the Wellcome Trust (WT105882MA). The funder played no role in the conceptualization, design, data collection, analysis, decision to publish, or preparation of the manuscript. M.S.S was funded in whole by Science for Africa Foundation to the Developing Excellence in Leadership, Training, and Science in Africa (DELTAS Africa) program [DEL-22-012] with support from Wellcome Trust and the UK Foreign, Commonwealth C Development Office and is part of the EDCPT2 programme supported by the European Union. For purposes of open access, the author has applied a CC BY public copyright license to any Author Accepted Manuscript version arising from this submission.

## Supplementary Materials

**Supplementary Figure 1.**
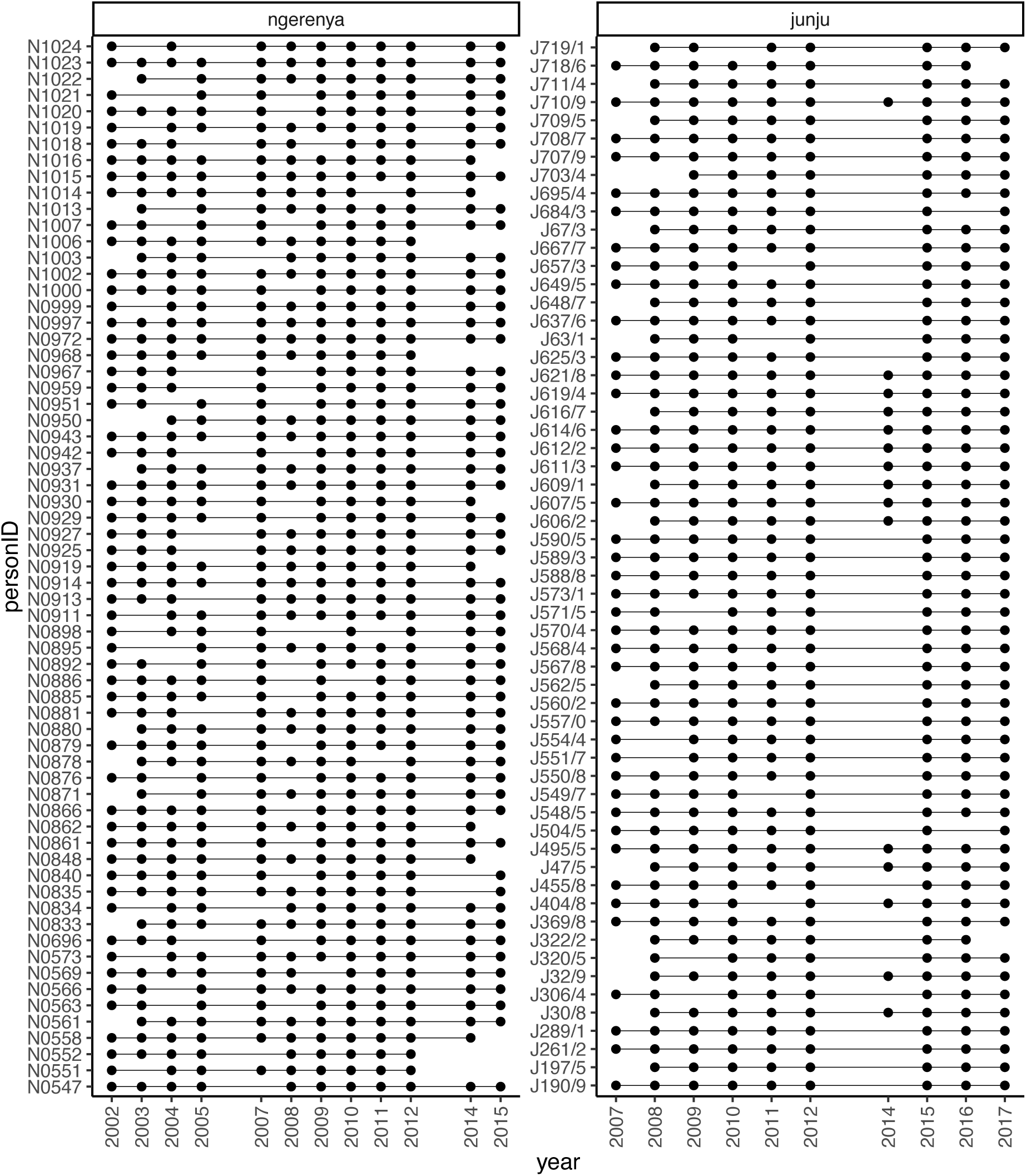
A summary of the sampling frame for the study cohorts (Junju and Ngerenya). Each vertical line represents the longitudinal time series for a single individual, and each dot represents a timepoint where a serum sample was collected.

**Supplementary Table 1.** Summary of pathogens, strains or subtypes, and corresponding antigens included in the immunoassay panel.

| Pathogen | Strain/subtype | Antigen |
| --- | --- | --- |
| Measles virus | Edmonston | Whole virus antigen |
| Coxsackie B virus | B1 | VP1 |
| Cytomegalovirus | TB40E | PP150 |
| Epstein-Barr virus | Type 1 | Nuclear antigen 1 |
| Herpes simplex virus | Type 1 | Extract |
| H1N1 Influenza A | A/California/07/2009 | Haemagglutinin |
| *Plasmodium falciparum* | 3D7 | AMA-1 |
| Rubella virus | HPV-77 | Whole virus antigen |

## Notes

### Competing Interest Statement

The authors have declared no competing interest.

### Summary of Updates

We have revised this preprint to address reviewer concerns and strengthen the robustness and interpretation of the findings. In particular, we addressed potential confounding through additional analyses, clarified methodological procedures, and expanded the Discussion to better reflect the limitations and interpretation of the findings.

